# Lived experiences of caregivers of persons with epilepsy attending an epilepsy clinic at a tertiary hospital, eastern Uganda: A phenomenological approach

**DOI:** 10.1101/2022.08.29.22279325

**Authors:** Lindah Okiah, Samuel Olowo, Stanely J. Iramiot, Rebecca Nekaka, Lydia VN. Ssenyonga

## Abstract

**Introduction:** Epilepsy has been found to affect caregivers’ quality of life, life style, psychological health, social well-being and their working time. Caregivers in Uganda as in the rest of the world are important in assisting a person with epilepsy in complying with medical directions and can be actively involved in communicating with health care professionals. Little is known about the lived experiences of caregivers of persons afflicted with epilepsy in Uganda. The purpose of the study was to determine the lived experiences of caregivers of persons with epilepsy attending the epilepsy clinic at Mbale regional referral hospital, eastern Uganda.

**Methods and materials:** Forty participants were selected for the study through purposive sampling. Face to face in-depth interviews with unstructured interview guide were conducted to gather participants’ information. The investigator conceptualized the interview guide, reviewed by co-investigators, and revised and approved as the final data collection instrument after an extensive and comprehensive literature review. The interview guide comprised of two sections, the first section comprised of the questions that elicited the participants’ social demographic information. The second section comprised questions that explored caregivers experiences of persons afflicted with epilepsy. Notations were taken and a digital recorder was used purposely for audio-recordings. All interviews lasted for an hour and were audio-recorded with the participants’ consent. An inductive thematic analysis was employed and adopted to identify the patterns emerging from the texts.

**Results:** The caregivers majorly perceived epilepsy as a burden. Four main themes were revealed from the analysis and these are; psychological, social, economic, and physical burdens.

**Conclusion:** The caregivers majorly perceived epilepsy as a serious burden. This burden can be psychological, social, economic, and physical. Therefore, services and plans targeting patients with epilepsy need to consider the burden that caregivers encounter to comprehensively manage epilepsy and its resultant burden.

## Introduction

Epilepsy remains a globally health challenge affecting all key health care players including caretakers [1]. The burden of refractory epilepsy is on the increase despite major pharmacologic advances in the management of epilepsy with over 22.5% of epileptic patients suffering from refractory epilepsy [1]. Studies have shown epilepsy to be common across the population, with devastating effects and increasing in prevalence rates [2]. The International League against epilepsy recognizes epilepsy as a chronic non-communicable disease and a significant public health challenge affecting over 70 million people worldwide [3]. Epilepsy accounted for 0.56% of the global burden of disease and therefore poses a major burden to the global health care community [4]. The disease burden from epilepsy has been reported to be higher in developing regions of the world [5]. According to a 2013 epidemiological report on sub-Saharan Africa, epilepsy was the fourth most common neurological condition among regions of Africa, with prevalence rates estimated to vary from 49 to 215 per 100,000 people [6]. The incidence of epilepsy has been reported to be high in Uganda with over 156 new cases of epilepsy per 100,000 people each year and an increasing prevalence of 13.3% [7]. Epilepsy affects everyone irrespective of race, culture, ethnicity, country, time and space [4]. Epilepsy and its resultant burden directly affect relationships between the caregiver and the care receiver [8]. It has a significant implication for the individual affecting the quality of life and indirectly affects the caregivers in various life domains [1]. The burden of epilepsy does not only arises from seizure activities but other factors like social and cultural stigma affiliated to epilepsy [9]. Social rejection of epilepsy victims, being barred form marriage and employment has also been associated with an increased burden of epilepsy in the community [10]. In East and South-central Africa, stigma from epilepsy has been associated with low school attendance, school attainment among children and increased inferiority complex among care givers in society [1]. Care giving and receiving for epilepsy can happen at any moment in life. “It involves a multidimensional role, varying from daily routine activities such as household management, checking and monitoring tasks, providing transportation and physical care to recognizing reportable symptoms” [11]. Care givers can further assist a person with epilepsy in complying with medical direction and can be actively involved in communicating with healthcare professionals [12]. The caregivers of persons with epilepsy experience severe emotional, physical, and economic burden resulting from the nature, chronicity, disability and stigma affiliated to epilepsy condition. Several studies globally have shown that substantial burden is impacted in various ways on caregivers who support individuals with epilepsy. While epilepsy is a common phenomenal in Uganda, there has been limited regional research done to study the lived experiences of care givers of persons with epilepsy. Moreover with reference to the Health Management and Information Systems, (HMIS) register at Mbale regional referral hospital psychiatric unit, the outpatient epileptic clinic enrolled a total number of 295 out of 831 new patients during the financial year July 2016-June 2017, 124 of the 295 persons afflicted with epilepsy were male clients and 171 were female clients, a total of 206 clients who returned for follow-up treatment were normally accompanied by their care givers. However, information concerning lived experiences of care givers of persons with epilepsy was insufficient. Therefore, this study aimed to determine the lived experiences of care givers of persons with epilepsy attending the epilepsy clinic at Mbale regional referral hospital, eastern Uganda.

## Materials and methods

### Study design

This study employed a qualitative phenomenological approach. A Phenomenology is an approach to qualitative enquiry that focuses on lived experiences of persons [13]. Through using this approach the research gains a profound understanding of the phenomenon. It therefore generates a detailed view of human experience.

### Study site and study population

The study was conducted at Mbale regional referral hospital Psychiatric Unit. Mbale regional referral hospital is located in Eastern Uganda approximately 214km East of Kampala, Uganda’s capital [14]. Mbale regional referral hospital receives patients of all age groups children, men and women who come from all over the 15 districts of the eastern region of the country [15]. It has a catchment population of 4 million, bed capacity of 511, bed occupancy averagely 359 with 18 inpatient wards [15]. The hospital has over forty physician including intern doctors, over 170 nurses and students (both medical and nursing) [16]. The psychiatric unit has an inpatient and runs an outpatients epileptic clinic every Wednesday, patients attending this clinic usually come along with their caregivers. The psychiatric unit has no trained medical doctor working in the unit [17]. The psychiatric unit is majorly covered by psychiatric clinical officers who hold diploma in clinical medicine and with a psychiatry specialization and psychiatric nurses with diplomas and certificates in psychiatry specialization [17]. The study population comprised of 30 adult care givers of epileptic patients who attended the epileptic clinic at Mbale regional referral hospital and 10 health workers who attended to patients with epilepsy at the hospital during the study period. This gives a total population of 40 participants for the study.

### Eligibility criteria

All accessible health workers and eligible caregivers who lived with the patients, were involved in monitoring treatment adherence and attended the follow up clinic with the patient were enrolled consecutively until the required sample size was obtained. Caregivers below 18 years, caregivers whose persons had other physical and mental disorders apart from epilepsy and those who refused to consent were excluded from the study.

### Sampling technique

Purposive sampling was employed to select only caretakers of persons living with epilepsy. For the sample size, saturation sampling was employed. Using this approach, saturation was assumed when no new information emerged. Full saturation was reached on a sample of 30 participants.

### Data collection

Face- to- face in-depth interviews with an unstructured interview guide were conducted to gather participants’ information. Unstructured interview [18] is a “conversation with a purpose” and enables researchers to obtain in-depth information. “It is also described as a shared experience in which researchers and interviewees come together to create a context of conversational intimacy in which participants feel comfortable telling their story”. The interview guide consisted of open-ended questions and follow-up probes to ensure a better understanding. The aim was to ascertain and explore lived experiences about care of a patient with epilepsy. The interview guide was pretested among caregivers attending the same clinic who consented to take part in the pilot study. Necessary adjustments were later done to ensure clarity. Results from the pilot survey were not considered during the final analysis. Before the interview began, verbal and written informed consent were obtained from the participants. Understanding was ensured among the prospective participant on the aims and scope of the study. The interview guide was conceptualized by the investigator, reviewed by co-investigators, and revised and approved as the final data collection instrument. This was after a thorough literature review. Distractions during the interview process was avoided as much as possible by conducting the interviews in quiet secluded rooms. The quiet rooms were assumed to be a neutral setting and would encourage freedom of speech among the participants. The presence of a trained and experienced qualitative researcher was ensured during the interview. The experienced qualitative researcher also assisted in taking note of the non-verbal cues. The interview lasted between 40minutes to 1hour. All caregivers were interviewed in the language which they best understood which were English, Lugisu or Luganda. The key informants (health workers) were interviewed in English. During interview guide preparations, a fruitful discussion was ensured for those English words that had no direct translation in the local languages and suitable ways of conveying them to participants were reached. Saturation was reached when no new information was emerging among the participants. The interviewers pre-briefed, had frequent reviews and debriefed each other throughout the interviews cycle. All what ensured was the preserve the original meaning in the participants’ perspective.

### Data analysis and presentation

The collection and analysis of the data were done simultaneously. The In-depth interviews were audio recorded and later subjected to a careful verbatim transcription. An inductive thematic analysis was employed and adopted to identify the patterns emerging from the texts. Flexibility was ensured to variations while noting regularities, contradictions and emerging patterns among participants’ views. A thorough reading and understanding of the transcripts was ensured to obtain an overall sense of the participants’ lived experiences. Line coding of the statements was employed to discover how the participants experienced the phenomenon. A thorough review of the transcripts were ensured and later a list of codes were developed. During the comprehensive review of the transcripts, attention was paid to the differences and similarities. The generated codes were later merged to form themes. The analysis proceeded till all the concepts were identified and the resultant categories were coherent and meaningful. Credibility was ensured by independent analysis of the data by the investigators and a trained and experienced qualitative researcher. On a later comparison of the analysis results, a high agreement was demonstrated. Conformability of the study is demonstrated by how the in empirical data is line with the study findings. Dependability was ensured by ensuring clarity in the research process. Data was presented narratively. The reporting of data was in accordance to consolidated criteria for reporting qualitative research (COREQ) guide lines [19].

### Ethical consideration

The proposal was presented to Busitema University faculty of health science scientific committee. The study was later approved by Cure Children’s hospital of Uganda Research and Ethics Committee and the **REC number is: CCHU-REC/08/019**. Participation in the study was voluntary and an informed consent was sought from all eligible caregivers and key informants. Before data collection, participants were given a detailed explanation of what the study was about then they were given a consent form to sign. All participants’ information was handled confidentially, all data were limited to the investigators and no name but rather study numbers of participants was used during and after the study. The study was voluntary, refusal or withdrawal from the study was addressed by explaining to the participants that the study was for academic purposes and that all information provided would be kept confidential. During data collection and data analysis individuals’ rights and consent was sought and assured of confidentiality and anonymity.

## Results

### Demographics of the caregivers

Out of the thirty care givers that were interviewed, 16(53%) were females and 14(47%) of the participants were males. Majority, 20(67%) of the participants were married, 09(30%) were single and 01(3%) of the participants was divorced. With regards to education, majority, 15(50%) of the participants never acquired formal education, 11(37%) acquired certificate level education, 03(10%) acquired diploma level in education and 01(3%) had degree-level education. With regards to tribes, majority, 20(67%) of the care givers were Bagishu, and the remaining 10(33%) of the participants were of different tribes. Professionally, 29(97%) of the participants comprised of security guards, farmers, students, businessmen and women, and 01(3%) of the participants was a nurse. About 10(33%) of the participants were biological fathers, 3(10%) of the participants were sisters to the patient, 13(43%) of the participants were biological mothers, 1(3%) was a spouse and 02(7%) of the participants were caregivers. About 13(43%) of the participants reported that their patients had epilepsy for more than 5 years, 08(27%) fell in the range between three to five years, 08(27%) of the participants fell in the range between one to two years and 10(3%) of the participants reported that their participants had suffered from epilepsy for less than a year.

### Burden of epilepsy

The caregivers perceived epilepsy as a major burden affecting different aspects of their lives: psychosocial, economic and physical aspect of their lives. This has been presented narratively below:

#### Psychological Burden

This is a situation where a caregiver was assessed with a tendency to get their stable mind, which is perceived as a threatening or excessive uneasiness. Psychological burden was discussed under; effects of caring for persons afflicted with epilepsy in day to day life, interference with personal relationship with others, time for one’s self, feelings after separation from the patients and behavioral influence.

#### Effects of caring for persons afflicted with epilepsy in day to day life

Psychological burden was expressed differently among care givers, concern about the future lives of their persons was conceptualized among the participants:

> *“I…. worry a lot that this child will die any time, I think a lot because all the time she gets sick even right now we are hospitalized here but when we are discharged I know she is going to get an attack at any time that makes me worry, my husband used to help me but when he realized that this disease cannot be cured he does not give me any help. I am the mother as well as the father to this child that disturbs me; I wish God could heal my daughter…”* [Care giver no.7].

> *“I… don’t know how my daughter will be in future; I wonder whether she will be fine…”* [Care giver no.12]

#### Effects of caring for a person afflicted with epilepsy on personal relationship

Feelings of being looked down upon or belittled for taking care of their loved one was expressed by the participants:

> *“… Some people fear me and they isolate themselves from me, some of them fear my son and think that he will infect their children nowadays I restrict him from moving to the neighbors’ homes, we stay in our home…”* [Care giver no.4].

However, some participants admitted that they were being well cared for:

> *“…Mmmh …we talk well with my friends and neighbors, there are some days when he gets an attack when I am not around but my friends and neighbors who like me come around and help me*…*”* [Care giver no. 17].

#### Effect of caregiving on individual’s time

Caregivers acknowledged that their time is really affected taking care of their loved ones:

> *“…Really I don’t have enough time because I have some other things that I would like to do but because of caring for the patient I feel that the time is not enough, as you know with business when you are not there you end up losing customers and making losses…”*[Care giver no.25].

> *“…I have enough time but when I go somewhere I make sure that I come back quickly because I know that any time my daughter can get the attack…”* [Care giver no.2].

#### Feelings after separation from their persons

Care givers had mixed feelings while they were separated from their loved ones; experiences of bad feelings when separated because of the close affection that they had developed with their person unfolded:

> *“…It affects me, I feel bad because I didn’t want him to be the way he is. You know…*.*in our hearts we are not the same there are good and bad people, I fear if I leave him behind he may be mistreated*…*”* [Care giver no.13].

However, a sense of hopefulness had not left others in that all was well while they were away from their persons:

> *“…I leave him with my daughter in-law she is good and she takes good care of him just like me…”* [Caregiver no.4].

Others expressed no worries at all because they had trust in God and the antiepileptic medication that they were using.

### Behavioral influence associated with care giving

This included positive or negative attitudes that the care givers developed while caring for their persons, a sense of good behavior still persisted among the participants. This was majorly affiliated to proper understanding of the patient’s condition resulting from past experience with the same patient:

> *“…Since I understand her condition, and I know that no one calls for a disease my behaviors have never changed in fact I treat her with a lot of care as a delicate person…”* [Care giver no.30].

Some behavioral changes surfaced among the participants:

> *“…For me normally my mood changes when he becomes resistant to what I feel he should do, I get annoyed with him there are times when he is supposed to return to the Hospital for review and he seems to have given up I use force to bring him to the hospital because when I talk softly to him he is never co-operative…”* [Care giver no. 16].

### Social Burden

This was measured basing on the understanding linked to outcomes such as; effects of epilepsy on public relationship, opinions about unpredictable epileptic attacks, feelings of stigma, alternative care givers in case of absence and effects of caregiving to persons living with epilepsy in a family:

#### Effect of caring for a person afflicted with epilepsy on public relationship

Feelings of bad public relations emerged among the participants:

> *“…That is so shameful I feel sorry because if she is in a public place and she makes noise, she begins fidgeting and eventually she falls down. Upon recovering, she gets up with a lot of energy like a mad woman and urinates on herself when this happens many people laugh at me and run away…. you just have to be strong …”* [Care giver no.7].

However, an intact public relations still persisted among some participants:

> *“…When she falls in a public place, some people run away, but those who know about this disease come and help me…”* [Care giver no.8]

#### Opinion about the unpredictable epileptic attacks

The participants were interviewed to seek their opinion about the unpredictable epileptic attacks. Concern about the situation stood out among the participants:

> *“I don’t feel okay because I am not prepared for these attacks, I feel a lot of pain!. if there were some signs showing me that my patient is going to get an attack I would make her sit or lie down in a safer place before it occurs so that she does not get injured…*.*”* [Caregiver no.11].

> *“…Obviously those attacks scare me utmost, it mainly scares me when I struggle to do what I am supposed to do and after doing it you see the thing coming again, at times I also feel stressed I ask myself if by now I am around and I am here what if she is alone in the room and it so happens that it comes then what could have happened…”* [Care giver no.14].

However, some expressed no feelings of worries because they were used to the situation.

#### Feelings of stigma

The participants were assessed on the basis of stigma that’s caused while caring for their loved ones. Experiences of discrimination in society while caring for their loved ones emerged among the participants:

> *“…Ok… some people chase him and tell him to come back home, some say that he will infect their children, I make sure that I restrict his movement, he stays at home, he eats, bathes, plays and sleeps thereafter…”* [Care giver no.4].

However, life remained normal among some participants:

> *“…Me I don’t mind about peoples words I just help him as a brother some people tend to isolate him but me I can’t, run away from him because they say blood is thicker than water I know the truth about this disease, I also believe that one day God will heal him*…” [Care giver no.20].

#### Alternative care givers in case absence

This situation was assessed on the basis of the care givers opinion in relationship to their absence from their persons in unavoidable circumstances. Fear of unknowns during their absence persisted among the participants:

> *“ …When I am not at home at least there has to be somebody who is close to him, I make sure that my phone is on so that I am notified if his condition changes, there I can call people who can help me to take him to the hospital…”* [Care giver no.20].

> *“…When I get a problem, I first take care of her, I prepare food for her and then I go away. When I am supposed to spend a night away from home, I excuse myself and tell them that I have a patient at home, and there is no one left behind to help her, so I get back home quickly and take care of my daughter…”*[Care giver no.8].

However, some caregivers expressed no worry because they have good substitute caregivers when they are away from home.

#### Effects of caring for persons afflicted with patients in a family

Sometimes families get affected while caring for their loved ones; however some families may not be affected with the situation. Negative effects expressed as ‘ unsettledness’ on their families as a result of caring for their loved ones emanated among the participants:

> *“…Men leave me because I can’t be there with them, when we want to have some good time together that’s when the child’s condition gets worse; in fact my daughter’s illness has made me fail to settle in marriage. All the men I get run away from me once they realize that she is suffering from that disease… “* [Care giver no.7].

> *“…For sure we feel desperate, we have tried our level best but things are not working out*…*”* [Care giver no.20].

However, some of the caregivers expressed no effect on their family.

### Economic Burden

This was defined in terms of the cost incurred towards medical care. Care givers are more likely to have economic burden. This was assessed basing on some of the factors such as; family economic interference, other challenges faced while caring for persons afflicted with epilepsy, effects of caring for a person afflicted with epilepsy on production at work, and the effects of care giving on financial stability:

#### Interference with the care givers source of income

Economic burden can affect both the patient and the care givers’ quality of life and access to medical care. These effects can be measured in a positive and negative way depending on severity of the disease. A significant impact on income sources was expressed emotionally by the participants:

> *“…There is less time to attend to business, no hope for employment opportunity due to stigma. At times when I get a job I end up losing it because of my brother’s sickness, I absentee myself from work to look after him, my income is just affected like that*…*”* [Care giver no. 25].

> *“…Caring for my daughter really affects me, I don’t work especially when she is sick, she needs much attention and when I don’t work being a tailor, I don’t get money, my customers get annoyed with me when their clothes are not ready, they quarrel at me and I end up losing many of them…”* [Care giver no.3].

#### Effects of caring for persons afflicted with epilepsy on production at work

Many care givers find it a rewarding task to look after their beloved ones, however for some people being a care giver has got a negative impact on their daily production at work. Different degrees of worries about productivity at work surfaced among the participants:

> *“…My productivity at work is partially affected, because the patient takes 50% of my time and I spend the other half for doing my work, I don’t give my best to my work and my family suffers the consequence…”* [Care giver no.30].
>
> *“…I have less time to attend to my business, I have lost customers and my earnings have reduced…”* [Care giver no.16].

#### Effects of caring for a person afflicted with epilepsy on financial stability

Depending on the severity of epilepsy, there may be other health needs which may be so costly. Financial instability was expressed by the participants in the most extreme form that is fears of potential resultant poverty:

> *“… Since I am spending much of my time caring for the patient with a chronic illness, I may end up becoming poor because I am not working but spending daily and I may remain without anything…”* [Care giver no.18].
>
> *“…If I spend much of my time caring for the patient, I will become poor because all I have will go to the patients feeding, transport, and drugs…”* [Care giver no. 10].

### Physical Burden

Physical burden constitutes the area of; physical burden to caring for a person afflicted with epilepsy, physical challenges that may affect the health of care givers, inconveniences associated with caring for persons afflicted with epilepsy at night and inconveniences of feeding one’s self while caring for a person afflicted with epilepsy:

#### Physical burden associated with caring for persons afflicted with epilepsy

According to the participants, some reported that it was difficult to care for their persons afflicted with epilepsy. Feelings of uneasiness during the care of the patients was expressed by the participants:

> *“…it is a little bit hard because an epileptic patient is not like any other patient when he develops an attack, you have to make sure that he doesn’t get an injury of which sometimes he has much energy and after he has developed the attack he gets sleepy, and confused so I have to carry him to the hospital it is a little hard but I just have to endure and move on…”* [Care giver no.2].
>
> *“…It is difficult because it’s a chronic illness that will take the rest of someone’s time so someone has to have enough time…”* [Care giver no.22].

#### Physical challenges that may affect the health of care givers for persons afflicted with epilepsy care

The physical burden faced by the care givers were based on the respondent’s opinion, some of the participants had challenges associated with giving care to their patients because they had to carry their patients to a safer place after a fit, they had to monitor their movement, and some of them had difficulties with feeding, especially after their persons fell:

> *“…*..*Yes at times I have to carry him and he is heavy, in this process I end up feeling chest pain, backache I really become tired especially when he collapses it is a big burden to me*…” [Care giver no.25].

> *“Yaah!. the headache is there because all the time I am thinking sometimes I feel as if I am getting ulcers…”* [Care giver no.16].

However, others reported that they were comfortable with the situation.

#### Inconveniences caused by caring for persons afflicted with epilepsy at night

Night inconveniences described as ‘ disrupted sleep’ and ‘no sleep’ evolved among the participants:

> *“ …The attacks mostly come at night, I have to wake up in the night and struggle to rush him to the hospital it is very frustrating especially when the attack comes in the midst of the night I feel like I am trapped and helpless…”* [Care giver no 20].

> *“My mind is ever pre-occupied with thoughts, I hardly get sleep it is worse when dates for review come and I don’t have money, since they also say that the disease is genetic I sometimes think that it may show up in other family member I just imagine how I will handle the situation if it so happens …”* [Care giver no. 16].

#### Burden of feeding while caring for a person afflicted with epilepsy

The burden here was not so often, this depended on the frequency of the attack and the severity of the disease. When the attack was frequent, the care givers feeding habits got affected and vice versa:

> *“…The way someone behaves makes you lose appetite for they can bring for you food but you can’t eat when she is in an attack…”* [Care giver no.22].

> *“…When she is attacked, I first wait for her to stabilize that is when I eat food if she does not stabilize my interest for food goes away…”* [Care giver no. 24].

However, others never lost appetite for food during the attack because they were used to the situation.

### Key informants

#### Findings

About 10 key informants were interviewed, 05(50%) of the participants were females and 05(50%) of the participants were male, 07(70%) of them were married whereas 03(30%) of the participants were single. With regards to education 07(70%) of the participants had attained Diploma-level education, 01(10%) of the participants was a degree holders, 01(10%) was a certificate holders and 1(10%) of the participants had studied up to senior four. Majority of the participants were of different tribes 6(60%), whereas 2(20%) of the participants were Itesot and about 02(20%) of the participants were Bagishu. Majority of the participants were Psychiatric nurses 06(60%), about 02(20%) of the participants were psychiatric clinical officers, 01(10%) of the participants was a counselor and 01(10%) of the participants was a peer support worker. Seven themes emerged out of the interviews that were carried out with the key informants.

#### Perception of the way care givers felt while taking care of their persons

Some of the participants thought that the care givers felt bewitched, cursed, and unfortunate. Some thought that the care givers developed bad feelings and they ended up giving up on the caregiving role and others thought that the care givers felt that health workers were knowledgeable and could therefore give them good advice and treatment:

> *“…I think they are stressed, they are also stigmatized because people have many beliefs about epilepsy in the community some people think that the patient got the disease because the family sinned or is cursed. These things lead to stress or stigma in the lives and families of the care givers…”* [Key informant no.5].

They also thought that the care givers felt that it was their obligation to care for their persons:

> *“…For them what they feel is that they have to be there for the client to ensure that they get the right drugs, at the right time, in the right dose, they also give a report about the patients progress to the health workers…*..*”* [Key informant no.7].

#### Institutional programs that dealt with the stress related effects of the care givers

Some of the participants acknowledged that the institution employed psychologists, psychiatric clinical officers, counselors, social workers, and psychiatric nurses among others who gave continuous health education. They counseled and encouraged caregivers not to give up on their role, some admitted that they had community outreach programs and radio talk shows where they sensitized the public about epilepsy, and one participant reported that they linked up the needy patient who come to the facility without care givers to the social workers for support:

> *“…We have outreach programs whereby we go out to the people with epilepsy in the community and talk to them about how they should take their medication, we tell them that epilepsy is a chronic illness and we encourage the care givers to support their persons, we also have radio programs whereby we go on air and talk about epilepsy and clear peoples misconceptions about this disease, we answer their questions and tell them about the services that are offered here at the hospital and we discourage them from seeking help from the traditional healers…*.*”* [Key informant no.5].

#### Role of health workers in caring for caregivers

Some of the participants reported that their role is to counsel patients, give health education talks, encourage drug compliance, protect the patients and their caregivers and encourage the care givers to continuously take care of their persons:

> *“…*.*My role is to teach them how to manage these patients, I tell them about the danger areas that they that they should protect their patients from like fire places, water bodies, or broken bottles because these things can cause harm to the patient…”* [Key informant no.2].

> *“…*..*My role is to help these care givers to appreciate that epilepsy is a life long illness and it is just like any other disease so taking care of a patient with this illness is just like taking care of a patient with any other illness. They have to appreciate that these fits can be controlled though it takes long and someone has to be on drugs throughout their life…*.*”* [Key informant no. 6].

#### Resources available in taking care of the epileptic patients and their care givers

Some of the participants reported that antiepileptic drugs were available, some said that the institution employed, trained health workers and others said that they received support from some non-government organizations, they had good security, and beds for the patients’ use:

> *“*……*We have specialized personnel, drugs are available and the mental health unit is fully functional it provides services to the patients 24 hours in a day……”* [Key informant no.3].

> *“…*..*We have antiepileptic drugs, patients provide meals for themselves but if there is one who does not have a relative, JENGA (A Christian based Non –Government-Organization) helps them to buy drugs, they provide food for them and at times they give the patients clothes. We also have social workers who take care of the patients who come to the hospital without their relatives…*.*”* [Key informant no.1].

#### Effects of caring for persons afflicted with epilepsy on family/social relationships

Some of the participants reported that they thought that families of the caregivers were stigmatized in the communities, and others thought that the care givers wasted resources, and lost their jobs and social network as well:

> *“…*..*I think that family relationships can be affected due to stigma, other people say that this thing is contagious they even fear to share personal effects, for instance if it is a child who is sick, the neighbors fear and stop their children from playing together with him or her. In my village I used to observe people serve these patients food on banana leaves they would refuse to use their plates thinking that this thing is contagious, this patients at times get irritable and if it is a man he can chase away the wife so their relatives can be disgusted with them…”* [Key informant no.1].

> *“…*..*Resources are wasted, money is wasted in caring for them, time is wasted in caring for them and they are stigmatized and think that the disease is contagious*…*”* [Key informant no.3].

#### Symptoms that care givers get as a result of caring for their loved ones

Some of the key informants reported that they thought that care givers got fatigued and the others thought that they presented with symptoms of stress, such as headache, worry, chest pain, and backache and sleep disruption:

> *“…*.*Sometimes they get fatigued because when the patients have a fit they can bite them when they are unconscious, they can also beat them because they become so energetic and in the process of restraining them they become tired…”* [Key informant no.6].

> *“…*.*I think they can develop headache, because of thinking so much about the patient’s condition that has no cure, also they can have disrupted sleep patterns because epilepsy attacks come even at night and interfere with their sleep…”* [Key respondent no.10].

#### Effects of care giving role on care givers personal well being

Some of the participants reported that they thought that the caregivers would end up becoming poor, and some reported that they thought that the caregivers experience social, physical and psychological burdens which manifested in form of productivity loss at work, loss of social network, loss of appetite for food, and disrupted sleep among others:

> *“…Most of them end up becoming poor because they are confused, people deceive them that traditional healers can solve their problem so they spend a lot of money and by the time they come to the hospital the patient’s condition may have deteriorated and they may not afford to buy the drugs that are needed to manage this severe form of epilepsy because they are very expensive…*.*”* [Key informant no.1].

> *“…*..*Because of stress they are ever thinking about how the patient will be at the end, they end up being lonely, psychologically tortured and live in segregation…”* [Key informant no. 2].

## Discussions

Epilepsy is a condition that is treatable but potentially devastating especially in low resource setting where access to treatment is not readily available [20]. Epilepsy does not only affect the primary bearer but also the caregivers [21]. This study therefore was to determine the lived experiences of care givers of persons with epilepsy attending the epilepsy clinic at Mbale regional referral hospital, eastern Uganda. The caregivers perceived epilepsy as a major burden interfering with different aspects of their lives.

Different psychological burdens surfaced among the caregivers. Worries about day-to-day living and future life of their persons was expressed by the participants and, disrupted or no sleep. The finding has been reported in other studies: a study in the United States [22] found that caregiving to patients with epilepsy adversely affects the caregiver’s psychological health with many caregivers reporting anxiety, depression and insomnia after becoming caregivers.

These findings is also similar with the findings of Settineri and friends [23] which showed that care givers rise in the level of anxiety. Studies in Nigeria [20] also found high levels of emotional distress among caregivers of patients with epilepsy. Caregivers find themselves in constant alertness situation; they mention being in a state of hyper vigilance and being anxious. These findings were also in line with the findings of other related studies [8, 24]. Psychological interventions therefore need to be targeted at care givers of people living with epilepsy.

A number of social burdens in the forms of stigma and discrimination were experienced by the caregivers; disrupted public relationship, feelings of shame, being under looked or despised because of caring or their persons afflicted with epilepsy emerged. Related studies in United States [25] reported stigma among caregivers of patients with intractable epilepsy. These study findings are in line with the findings of other studies [23, 26-28] where different social consequences of epilepsy on the caregivers were reported. Many of these social consequences remain among the unmet needs in epilepsy and therefore further significantly contributes to the burden of epilepsy among the population [29].

This study showed that care givers of persons afflicted with epilepsy had more economic burden since they had to spend most their time caring for their persons this impacted negatively in their daily functioning. This finding was in line with that of other studies [30] which showed that epilepsy was noted to have significant economic implications in terms of health care needs, premature death and lost work productivity. This study revealed that caring for persons afflicted with epilepsy had a negative impact on their earnings as most of the time care givers had to be close to their persons than to their business, this was in line with the findings of other studies [1] which revealed that house hold members spend time and resources supporting the epileptic or paying for disability related costs increases the likelihood of the caregiver becoming poor. This study finding further agreed with the findings of [31] which showed that caregivers gave account of spending money to care, losing some days of work for care giving, and travelling to seek care for relatives. Absenteeism was associated with care givers leading to loss of jobs in some instances. High costs resulting from care of persons with epilepsy which occur either directly or indirectly has been reported in other related studies [22, 31, 32]. It’s therefore important to consider the economic component when understanding the burden of epilepsy.

This study found out that caregivers experience various physical burden; fatigue, chest pain and headache. This finding agree with the findings of other related studies [10] which showed that family care giver symptoms such as, fatigue, headaches, joint and muscle pains. Related studies in the United States [22] also found physical burdens like heads as quite commonly experienced by the care givers. Patients with frequent seizures was mentioned to more likely to be of higher burden to the caregivers. This is due to the fact that they require energy to carry them from places of attack to safer places. This has been reported in other studies elsewhere [22].

Majority of the key informants reported that they administer treatment to the patients, they offer counseling services to patients, and they give continuous health education talks at the facility and conduct radio talk shows. However, findings from the central part of Uganda among the epilepsy patients at Mulago national referral hospital found limited patient-health worker interaction [33]. They admitted having enough stocks of antiepileptic drugs and an adequate number of staff, they also reported that some of the patients and their caregivers were receiving support from the social workers and some Non-Governmental Organizations. Majority of the key informants believed that caregivers are stigmatized in the communities, some of them reported that the care givers wasted resources (money and time) while caring for their persons and had lost their jobs and social network. They also thought that caregivers got symptoms like fatigue, stress, headache and sleep disruptions these findings matched with the findings of most of the studies done in other parts of the world [26, 27].

## Conclusion

This study looked at the lived experiences of care givers of persons afflicted with epilepsy attending the epilepsy clinic at Mbale regional referral hospital and revealed that care givers. The caregivers majorly describe epilepsy as a major burden affecting different aspects of their lives. Therefore, findings of this study revealed that the care givers of persons afflicted with epilepsy experience psychological, social, economic, and physical burdens as they make efforts to care for their loved ones.

## Recommendations

The health care providers should continue targeting care givers of persons afflicted with epilepsy in their service provision to ensure that their state of health is not adversely affected. Services and plans targeting patients with epilepsy need to put into consideration the burden that caregivers encounter in order to comprehensively manage epilepsy and its resultant burden.

## Data Availability

The data will only be available to individuals upon request for non-financial utilization purposes only. Individuals who have financial intentions will not be allowed access to the data

## Declarations

### Ethical approval and consent to participate

This study was approved by Cure Children’s hospital of Uganda Research and Ethics Committee and the **RCE number** is: **CCHU-REC/08/019**. Participation in the study was voluntary and an informed consent was sought from all eligible caregivers and key informants.

### Consent for publication

Not applicable.

## Availability of data and materials

Data from this study will be made available by the corresponding authors on a reasonable request.

## Funding

No funding was received towards this study however the researchers do acknowledge support Busitema University Directorate of Graduate Studies, Research and Innovation (DGSRI) for small grants programs that facilitated the writing of this manuscript.

## Acknowledgement

We are greatful to the study participants.

## Authors’ contributions

LO, SO and LVS conceptualized the study; LO, SO, LVS, IJS, RN, formal analysis and interpretation of the data; LO, SO, LVS and IJS wrote the original manuscript draft; LO, SO, LVS, IJS and RN performed review and editing of the manuscript draft. All authors read and approved the final manuscript.

## Notes

### Competing Interest Statement

The authors have declared no competing interest.

### Funding Statement

The author(s) received no specific funding for this work.

### Author Declarations

The study was approved by Cure Children’s hospital of Uganda Research and Ethics Committee and the REC number is: CCHU-REC/08/019. P.O. Box 903, Mbale, Uganda. Tel:+256 454 435 273

